# “I Don’t Do What I Learn”: A Study of Diet and Quality of Life Among Medical Students

**DOI:** 10.64898/2025.12.02.25341511

**Authors:** Pâmella Costela, Mariana Salles Kehl, Alexandre Sizilio, Renata Kobayasi, Rosimeire Queiroz, Andred Meirelles, Gustavo Nogueira Nóbrega, Mariana Toledo, Daniel Trindade, Marcelo Arruda Cândido, Patricia Tempski, Mílton A. Martins

**Affiliations:** Center for the Development of Medical Education, University of São Paulo Medical School, São Paulo (SP), Brazil

**Keywords:** medical students, ultra-processed foods, quality of life, sleep, physical activity

## Abstract

**Background:** Medical students are exposed to behavioral risk factors that may compromise health and well-being. We examined dietary patterns –with emphasis on ultra-processed food consumption – and their associations with quality of life, sleep, and physical activity in Brazilian medical undergraduates.

**Methods:** In this cross-sectional study, 105 students from a large public medical school completed validated assessments of diet quality, quality of life, sleep, and physical activity. Anthropometry and bioimpedance were obtained under standardized procedures. Group differences by diet-quality categories were tested with appropriate parametric or non-parametric methods. The significance threshold was two-sided α = 0.05.

**Results:** Participants had a mean age of 22.2 ± 3.4 years (52.4% male, 44.8% female, 2.9% other/undeclared). Diet quality classified 31.4% with ≤31 points, 37.1% with 31–41, and 31.4% with >41. Healthier diet profiles were associated with higher quality-of-life scores in Psychological (67.9 ± 13.9 vs 59.0 ± 13.4 and 58.8 ± 15.2; p = 0.012), Social relationships (77.0 ± 18.6 vs 61.8 ± 22.5 and 67.2 ± 17.7; p = 0.006), and Environment (72.0 ± 13.0 vs 61.7 ± 15.1 and 62.2 ± 16.5; p = 0.008) domains, and with better sleep (Pittsburgh Sleep Quality Index 5.6 ± 2.1 vs 7.3 ± 2.9 and 6.6 ± 3.4; p = 0.050). Physical activity increased across diet-quality strata (vigorous activity 3481.2 ± 3123.6 vs 1925.1 ± 2655.9 and 1230.3 ± 2091.3 metabolic-equivalent minutes/week; p = 0.003; total activity 4734.0 ± 3193.9 vs 3005.2 ± 2987.3 and 2184.6 ± 2186.2; p = 0.001). There were no differences across diet groups for depression, anxiety, or stress subscales, nor for bioimpedance indices (all p ≥ 0.109). Sex differences appeared only in body composition (greater skeletal muscle and lower percent body fat in males; both p < 0.001), with no sex differences in quality of life, sleep, or physical activity.

**Conclusions:** Among medical students, dietary patterns – particularly lower consumption of ultra-processed foods – were meaningfully related to better quality of life and sleep, and to higher physical activity. These findings can inform campus-level health promotion and student support strategies.

## Introduction

Human eating behavior is a multifaceted phenomenon encompassing biological, social, cultural, symbolic, and historical dimensions. Over recent decades, profound changes in dietary practices have been observed worldwide, primarily driven by the industrialization and globalization of food systems. The gradual replacement of fresh and minimally processed foods with ultra-processed foods (UPFs) has characterized the so-called “nutritional transition,” carrying deleterious implications for public health and population quality of life.

Ultra-processed foods are industrial formulations containing little to no whole food in their composition. They predominantly consist of ingredients extracted from foods, along with colorants, flavor enhancers, emulsifiers, substances intended exclusively for industrial use, and other artificial additives designed to modify sensory properties^1^. Although highly palatable, affordable, and shelf-stable, these products are nutritionally unbalanced and have been linked to increased prevalence of obesity, type 2 diabetes, cardiovascular diseases, certain cancers, and mental health disorders such as anxiety and depression.

The NOVA classification^2^ transformed the understanding of how industrial food processing impacts human health by designating UPFs as a central driver in the deterioration of contemporary dietary patterns. Unlike fresh or minimally processed foods, UPFs are engineered to be hyper-palatable, convenient, and commercially profitable, while lacking nutritional value and being closely associated with reduced food autonomy, rising rates of non-communicable chronic diseases, and compromised physical and psychological well-being. According to these authors, the global food system has increasingly been shaped by products that displace traditional culinary practices and promote a consumption logic rooted in artificiality and market appeal, with far-reaching consequences for lifestyles and collective health.

Brazil is no exception to this trend. Data from the VIGITEL Survey^3^ and demonstrate a marked increase in UPF consumption, often at the expense of fresh foods and home-prepared meals, particularly among urban populations and vulnerable groups such as students. Within this context, medical students represent a population of particular interest. This group frequently reports inadequate dietary habits, low engagement in physical activity, sleep deprivation, and diminished subjective well-being. Paradoxically, these same individuals – future healthcare providers, frequently fail to integrate into their own lives the very principles of healthy living that they will be expected to advocate professionally.

Our primary objective was to investigate the relationship between dietary patterns (with particular emphasis on UPF consumption) and various dimensions of health and quality of life among medical students. Our guiding hypothesis was that students exhibiting healthier eating practices, particularly characterized by lower UPF consumption, would demonstrate better quality of life, physical health, and psychological well-being.

## Materials and methods

### Study Design

This is a cross-sectional study conducted as part of the Project “Health and Quality of Life of Brazilian Medical Students and Their Relationship with Professional Training”, funded by the São Paulo Research Foundation (FAPESP; Grant No. 2019/13850-9) and approved by the Research Ethics Committee of the University of São Paulo Medical School (CAAE 56043522.9.0000.0068). All students signed a written informed consent form.

### Participants

Eligible participants were medical students of both sexes, aged 18 years or older, who were formally enrolled in the undergraduate medical course of the School of Medicine of the University of São Paulo. Exclusion criteria were students who did not complete all data collection procedures. Sampling was non-probabilistic and convenience-based. Data for this first analysis were collected between 10 January 2022 and 30 June 2024. Participants were recruited by members of the research group through posters displayed across the university campus, personal and/or institutional dissemination (e.g., in classes, departmental communications), and direct contact with the research team, as well as via peer referral, whereby students who had already enrolled in the study were encouraged to invite their classmates to participate.

### Data collection

Participants attended in-person sessions at our research laboratory, scheduled by prior appointment. After reading and signing the informed consent form, participants completed validated psychometric instruments and underwent anthropometric assessments and objective health measurements. Data collection was conducted by a trained team.

## Instruments and Variables

### Dietary Habits

Dietary patterns were assessed using the validated questionnaire^6^ “How is your diet?”, composed of 24 items rated on a four-point Likert scale (ranging from “never” to “always”). The scale measures four dimensions: (1) meal planning, (2) household organization, (3) eating behaviors, and (4) food choices. The total score ranges from 0 to 72 and classifies participants into three qualitative categories: low (<32 points), intermediate (between 32 and 41 points), and high dietary quality (more than 41 points).

### Body Composition

Body composition was measured via bioelectrical impedance analysis using the InBody 120® device (multifrequency, segmental, tetrapolar), providing estimates of body weight, lean mass, fat mass, and body fat percentage.

### Height

Height was measured with a digital stadiometer (HM-210D model) under standardized conditions (upright posture, barefoot, without head adornments).

### Physical Activity

Physical activity was evaluated using the International Physical Activity Questionnaire -Short Form (IPAQ-SF): a self-reported instrument that collects data on the frequency, duration, and intensity of daily physical activities. This questionnaire was translated and validated to Brazilian Portuguese^22^.

### Sleep Quality

Sleep was assessed using the Pittsburgh Sleep Quality Index (PSQI-BR), a 19-item instrument evaluating seven domains; total scores range from 0 to 21, with higher scores indicating poorer sleep quality. This questionnaire was translated and validated to Brazilian Portuguese^20^.

### Quality of Life

Quality of life was evaluated using the WHOQOL-BREF questionnaire, that consists of 26 items clustered in four domains: environment, psychological, social relationships and physical health. Answers are given on a 5-point Likert scale and points within each domain are linearly transformed to a score from 0 to 100, and higher scores represent better QoL. This questionnaire was translated and validated to Brazilian Portuguese^23^.

### Mental Health

Mental health symptoms were evaluated using the Depression, Anxiety, and Stress Scale (DASS-21), a 21-item scale comprising three subscales for depression, anxiety, and stress. This questionnaire was translated and validated to Brazilian Portuguese^24^.

### Data Analysis

For the statistical analysis we used the software SPSS (Statistical Package for Social Sciences) version 22. The level of significance was established at P < 0.05. To compare values of men and women we used unpaired T test. For comparisons among the three group of medical students according to the categories of the nutrition questionnaire, we used one-way analysis of variance followed by Bonferroni test.

## Results

The sample comprised 105 medical students, with a mean age of 22.2 years (SD = 3.4; range: 17–36), of whom 52.4% identified themselves as males (n = 55), 44.8% as females (n = 47), and 2.9% (n = 3) identified with another gender or chose not to disclose. Regarding their stage of training, 50.5% of participants were in the first two years of the program (basic cycle), 39% in the third or fourth years (clinical cycle), and 10.5% in the final two years (medical internship/clinical clerkship).

In terms of dietary patterns, 83.8% self-identified as omnivores, while 10.5% reported following an ovolactovegetarian diet, 1% an ovo-vegetarian diet, and 3.8% adhered to a strict vegetarian diet. Regarding dietary quality, as assessed by the “How is your diet?” questionnaire, 31.4% (n = 33) scored ≤31 points; 37.1% (n = 39) were in the intermediate range; and 31.4% (n = 33) scored >41 points, indicative of a healthy dietary pattern.

Table one shows the values of WHOQOl-bref, DASS-21, PSQI and IPAQ questionnaires, comparing male and female medical students. IPAQ results were transformed in MET equivalents (metabolic equivalents of task). We did not observe significant differences when male and female values were compared. Regarding mental health, DASS-21 scores revealed low-to-moderate levels of depression (M = 4.7; SD = 4.7), anxiety (M = 3.3; SD = 3.5), and stress (M = 7.4; SD = 4.4). Considering bioimpedance values, as expected, males had more muscle mass and less body fat than females.

**Table 1.**
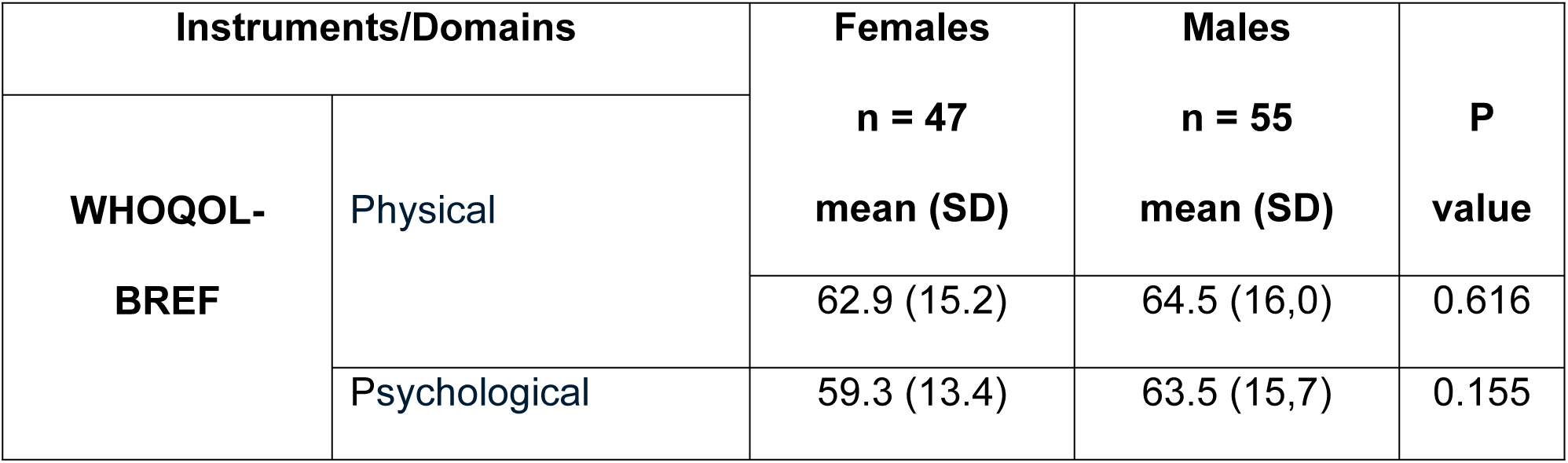
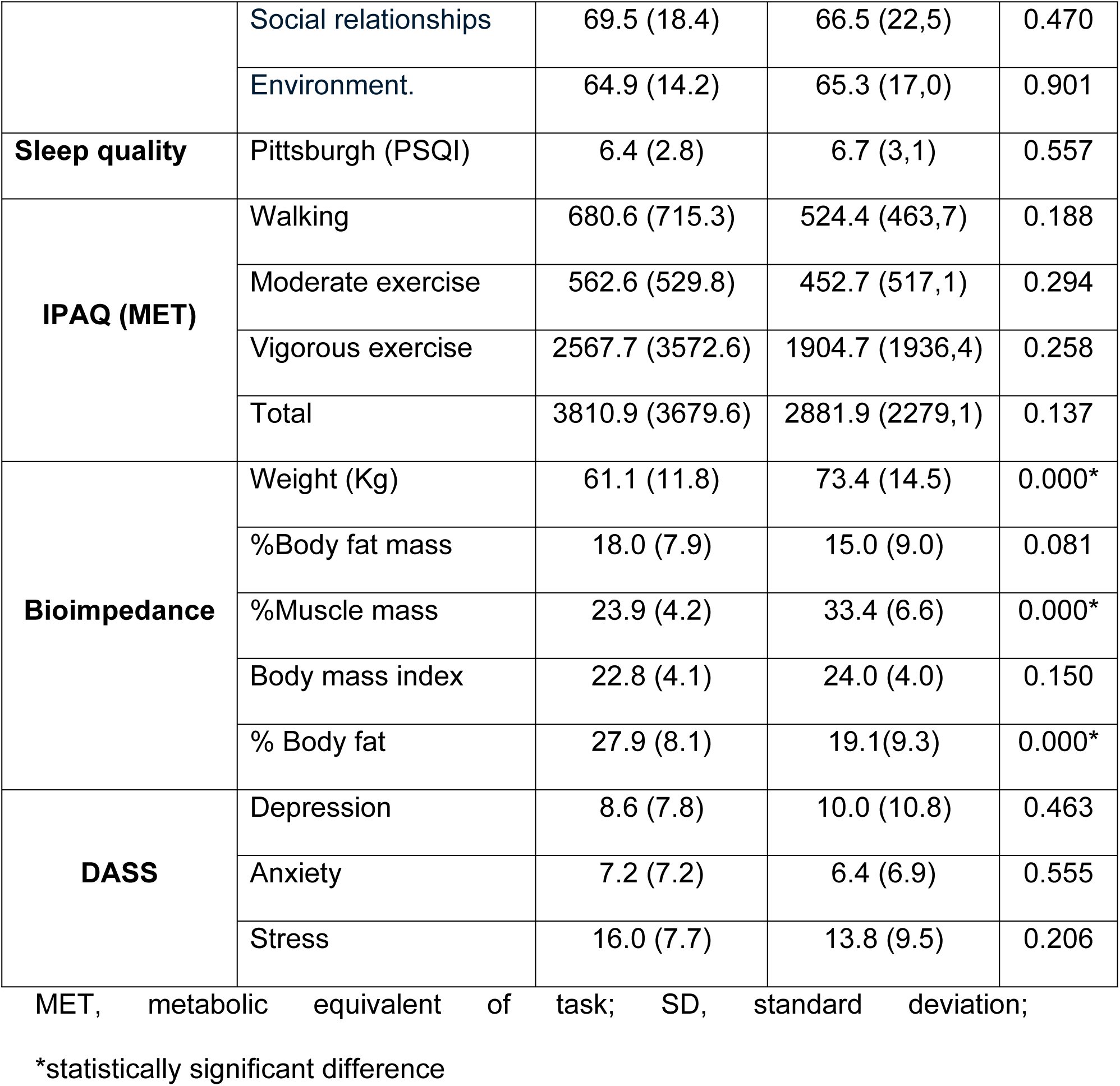
Results of the questionnaires and bioimpedance values

Table 2 shows the comparisons of the results of the three groups of students considering the results of the questionnaire of nutritional habits. Medical students with better nutritional habits showed significantly higher scores of three domains of the WHOQOL-bref questionnaire (Psychological, Social relationships and Environment). In addition, they tended to have a better quality of sleep (p = 0.05).

**Table 2.**
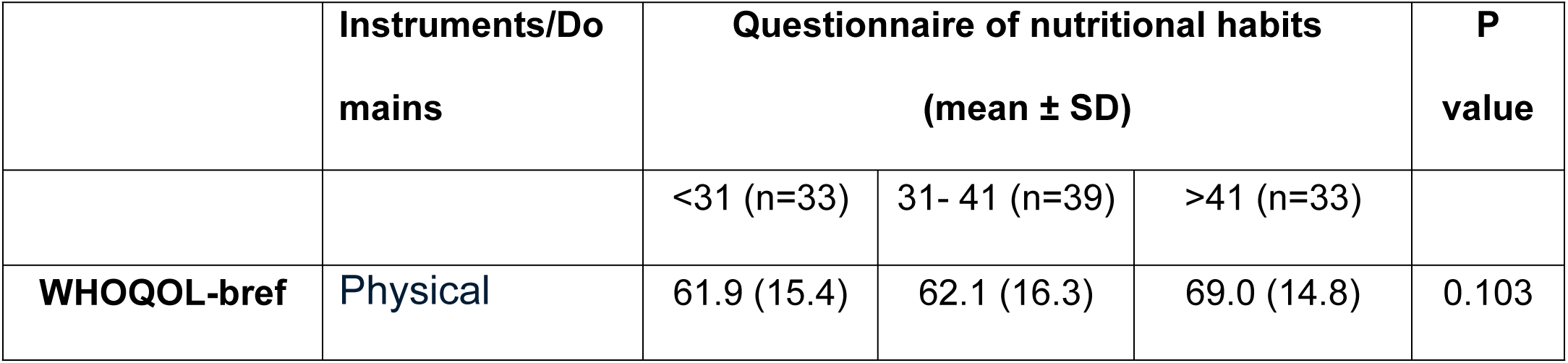
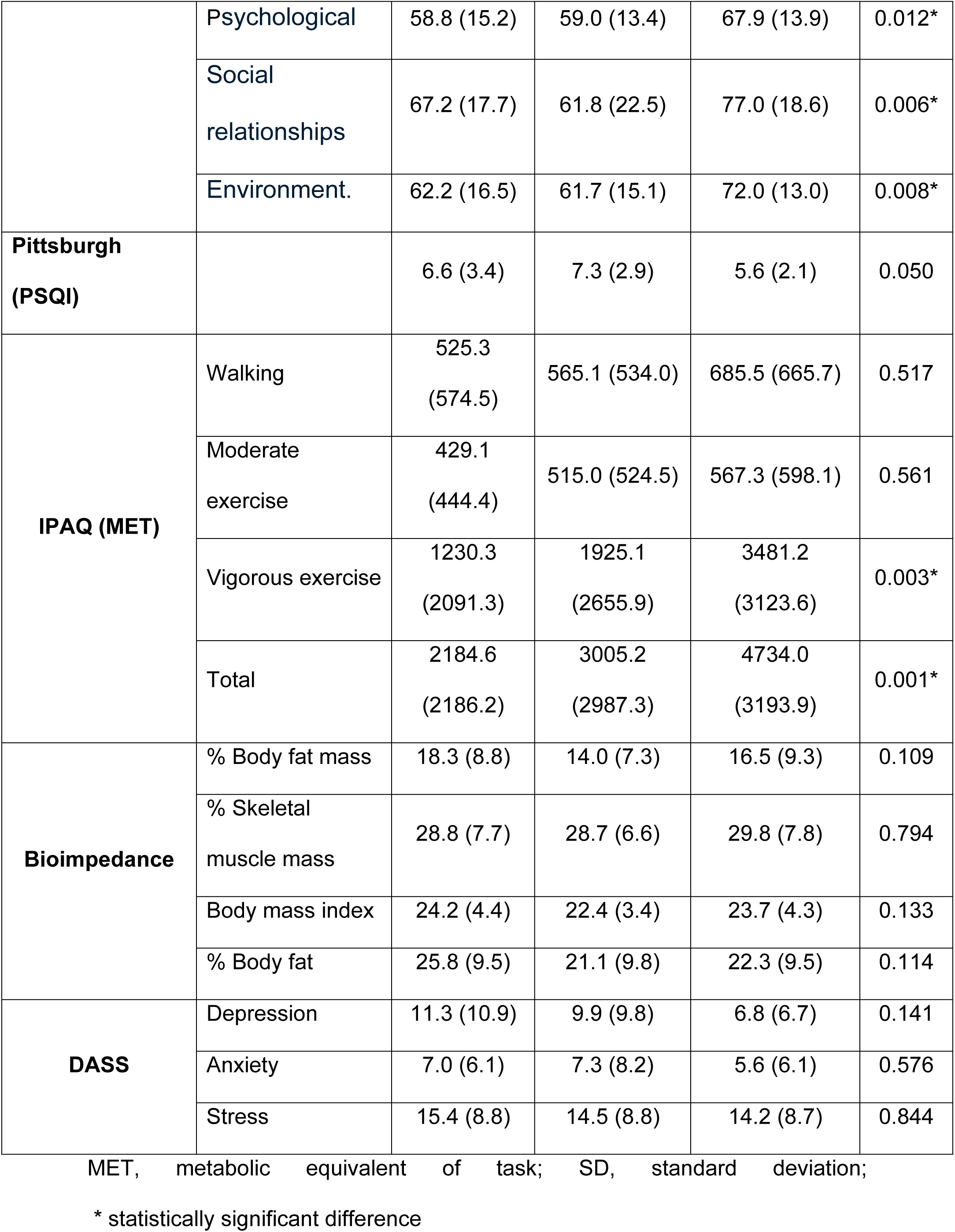
Comparisons of the three groups of medical students according to the results of the questionnaire of nutritional habits

| | Instruments/Do<br>mains | Questionnaire of nutritional habits<br>(mean ± SD) | | | P<br>value |
| --- | --- | --- | --- | --- | --- |
|  |  | <31 (n=33) | 31- 41 (n=39) | >41 (n=33) |  |
| **WHOQOL-bref** | **Physical** | 61.9 (15.4) | 62.1 (16.3) | 69.0 (14.8) | 0.103 |
|  | Psychological | 58.8 (15.2) | 59.0 (13.4) | 67.9 (13.9) | 0.012* |
|  | Social relationships | 67.2 (17.7) | 61.8 (22.5) | 77.0 (18.6) | 0.006* |
|  | Environment. | 62.2 (16.5) | 61.7 (15.1) | 72.0 (13.0) | 0.008* |
| **Pittsburgh (PSQI)** |  | 6.6 (3.4) | 7.3 (2.9) | 5.6 (2.1) | 0.050 |
| **IPAQ (MET)** | Walking | 525.3 (574.5) | 565.1 (534.0) | 685.5 (665.7) | 0.517 |
|  | Moderate exercise | 429.1 (444.4) | 515.0 (524.5) | 567.3 (598.1) | 0.561 |
|  | Vigorous exercise | 1230.3 (2091.3) | 1925.1 (2655.9) | 3481.2 (3123.6) | 0.003* |
|  | Total | 2184.6 (2186.2) | 3005.2 (2987.3) | 4734.0 (3193.9) | 0.001* |
| **Bioimpedance** | % Body fat mass | 18.3 (8.8) | 14.0 (7.3) | 16.5 (9.3) | 0.109 |
|  | % Skeletal muscle mass | 28.8 (7.7) | 28.7 (6.6) | 29.8 (7.8) | 0.794 |
|  | Body mass index | 24.2 (4.4) | 22.4 (3.4) | 23.7 (4.3) | 0.133 |
|  | % Body fat | 25.8 (9.5) | 21.1 (9.8) | 22.3 (9.5) | 0.114 |
| **DASS** | Depression | 11.3 (10.9) | 9.9 (9.8) | 6.8 (6.7) | 0.141 |
|  | Anxiety | 7.0 (6.1) | 7.3 (8.2) | 5.6 (6.1) | 0.576 |
|  | Stress | 15.4 (8.8) | 14.5 (8.8) | 14.2 (8.7) | 0.844 |

MET, metabolic equivalent of task; SD, standard deviation;
\* statistically significant difference

Students with healthier eating patterns also exhibited higher physical activity with higher overall and vigorous exercise IPAQ scores. We did not observe significant differences concerning mental health and bioimpedance scores.

## Discussion

The findings of this study indicate an association between healthier eating practices -particularly in relation to household organization, meal planning, eating behaviours, and food choices, including reduced intake of ultra-processed foods (UPFs), and higher levels of quality of life and physical activity among medical students. These results are consistent with the “golden rule” of the *Dietary Guidelines for the Brazilian Population*, which advises: “*always prefer fresh or minimally processed foods and home-prepared meals over ultra-processed foods*”^4^, as well as with international evidence identifying UPFs as major risk factors not only for physical health outcomes, such as obesity and metabolic disorders, but also for adverse effects on mental health and psychosocial functioning^5–8^.

Contemporary literature increasingly highlights that higher UPF consumption is linked to greater prevalence of depressive symptoms, anxiety, fatigue, and poorer academic performance^9^. These effects are hypothesized to be mediated through mechanisms such as low-grade systemic inflammation, gut microbiota dysbiosis, and neurochemical alterations affecting satiety and reward pathways^10–11^. In our sample, students with higher scores on the “How is your diet?” questionnaire also exhibited higher levels of physical activity, better sleep efficiency, and elevated scores in the physical and psychological domains of the WHOQOL-BREF and VERAS-Q, suggesting greater self-regulation and engagement in health-promoting self-care behaviors.

To increase the proportion of adults who practice physical activity regularly, it is very important that medical students, residents and physicians in general provide adequate counselling to their patients. It has been shown in medical students that personal physical activity levels are correlated with the frequency of physical activity counselling of their patients^12–16^. In addition, it has been shown that physicians and medical students with a normal body mass index (BMI) and who practice moderate and/or vigorous physical activity are more likely to feel confident about counselling their patients about physical activity than their colleagues who do not practice physical activity or are overweight^17^. It is important that counselling about the practice of physical activity also includes its impact on QoL. However, there are few data concerning the relationship between QoL and physical activity in medical students and physicians.

Medical students are often immersed in clinical and care-related demands, affording them less flexibility for self-care and structured eating routines. This trend diverges from prior studies reporting progressive improvement in health behaviors throughout training, culminating in more conscious choices during the final years^18–19^. Such discrepancies underscore the importance of considering curricular and institutional specificities, as well as the cumulative effects of stress and workload in advanced stages of medical education.

Regarding sleep, mean scores from both the PSQI; validated for the Brazilian population^20^ suggested insufficient or suboptimal patterns, although largely within non-pathological limits. Sleep deprivation among medical students is well-documented^21^ and is associated, as observed here, with poorer quality of life and higher prevalence of stress and anxiety symptoms. These findings suggest that healthy eating behaviors may act as a protective factor, mitigating the deleterious impact of sleep deprivation and academic demands on students’ overall health.

We previously evaluated a random sample of 1350 medical students from 22 medical schools and compared the volume of leisure time physical activity and quality of life. Forty per cent of the medical students reported no leisure time physical activity (46.0% of females and 32.3% of males). In contrast, 27.2% were classified in the group of high volume of physical activity (21.0% of females and 34.2% of males). Using the group that reported no leisure time physical activity as the reference group, we observed a strong dose-effect relationship between the volume of leisure time physical activity and quality of life in both male and female medical students.

Importantly, this study is innovative in integrating multiple dimensions of student life – including nutritional status, body composition, mental health, sleep, and physical activity, into a comprehensive model aimed at examining the influence of UPFs on medical students’ quality of life. This approach, grounded in validated instruments, provides a foundation for the development of educational and institutional interventions that foster a healthier, more critical, and sustainable learning environment.

## Conclusions

The findings presented herein show an association between healthier dietary practices -particularly those characterized by lower consumption of ultra-processed foods (UPFs), and improved indicators of physical and mental health, as well as higher quality of life among medical students. Participants who reported reduced UPF intake also exhibited greater engagement in physical activity, better sleep quality, higher levels of subjective well-being, and lower prevalence of stress, anxiety, and body dissatisfaction.

These results underscore the centrality of diet as a key determinant of overall health and a marker of self-care, particularly in populations vulnerable to psychological distress and self-neglect, such as medical students. Within the demanding context of medical education (marked by academic overload, performance pressure, and limited time) dietary patterns emerge as a privileged indicator of student well-being and their ability to regulate daily rhythms.

Accordingly, this study provides relevant empirical evidence to inform institutional strategies aimed at fostering healthier educational environments. Such strategies may include improving access to high-quality food on university campuses, creating designated spaces for rest and recuperation, and incorporating pedagogical initiatives that promote self-care, active listening, and well-being among future healthcare professionals.

Further research is warranted, particularly through larger samples and longitudinal designs, to assess the impact of targeted nutritional and behavioral interventions throughout medical training. The integration of institutional health policies with the ethical and technical training of students may contribute to the development of healthier physicians – professionals who are more attuned to their own limits and better equipped to promote comprehensive patient care.

## Data Availability

The anonymized dataset underlying the findings of this study will be made fully available in a public, open-access repository upon acceptance and publication of the article.

